# Platelet size as a predictor for severity and mortality in COVID-19 patients: a systematic review and meta-analysis

**DOI:** 10.1101/2021.07.15.21260576

**Authors:** Sarah Daniels, Hua Wei, David W. Denning

**Affiliations:** Division of Population Health, Health Services Research & Primary Care, School of Health Sciences, University of Manchester, Manchester, England; Division of Infection, Immunity & Respiratory Medicine, School of Biological Sciences, University of Manchester, Manchester, England; Manchester Academic Health Science Centre, University of Manchester, Manchester, England

**Author notes:** Correspondence: Sarah Daniels, The University of Manchester, Ellen Wilkinson Building, Oxford Road, Manchester, M13 9PL.

**Keywords:** COVID-19, mean platelet volume, platelet distribution width, platelet-large cell ratio, thrombosis

## Abstract

**Background:** Parameters reflecting platelet size can be sensitive indicators that circulating platelets are activated and COVID-19 patients are at increased risk of thrombosis. This systematic review aims to assess the association of mean platelet volume (MPV), platelet distribution width (PDW) and platelet-large cell ratio (P-LCR) with disease severity and mortality in COVID-19 patients.

**Methods:** English and Chinese databases were searched electronically to identify studies reporting data on MPV, PDW or P-LCR in COVID-19 patients. Included articles underwent a quality rating. A meta-analysis was performed using the standard mean difference and interpreted as the common language effect size (CLES).

**Results:** Twenty-two studies (11,906 patients) were included in the meta-analysis. Of these, 14 were rated poor and eight were fair. The MPV and P-LCR was significantly higher at hospital admission in severe patients compared to non-severe patients. The MPV, PDW and P-LCR were significantly higher at hospital admission in non-survivors compared to survivors. There was a marked increase in the probability of a severe COVID-19 patient presenting with higher P-LCR at hospital admission than a non-severe patient (CLES: 68.7% [95% CI: 59.8%, 76.5%]), when compared with MPV and PDW ((CLES: 59.2% [95% CI: 53.1%, 65.1%]) and (CLES: 55.9% [95% CI: 50.6%, 62.2%]), respectively).

**Conclusion:** Severe COVID-19 disease is associated with the increased production of larger, younger platelets. When comparing MPV, PDW and P-LCR, P-LCR is the most important biomarker for evaluating platelet activity. P-LCR testing at hospital admission could identify COVID-19 patients with increased risk for thrombotic events, allowing preventative treatment.

**Summary Table:** *What is known on this topic:*

- The incidence of thrombotic complications is high in COVID-19 patients with severe disease.
- Parameters reflecting platelet size can be sensitive indicators that circulating platelets are activated and that COVID-19 patients are at increased risk of thrombosis.

*What does this paper add:*

- When compared to MPV and PDW, P-LCR is the most important biomarker for evaluating platelet activity in COVID-19 patients at hospital admission and could be used to identify patients with increased risk for thrombotic events.
- Current evidence is predominantly derived from retrospective design. Prospective studies are warranted to accurately determine cut-off values that may be used in the clinical setting.

## BACKGROUND

Coronavirus disease 2019 (COVID-19) disease is caused by severe acute respiratory syndrome coronavirus 2 (SARS-CoV-2). While most cases of COVID-19 are mild, some develop severe viral pneumonia with respiratory failure, that can result in death. Severe disease is predominantly observed in the elderly and those with underlying health conditions such as hypertension, diabetes and coronary heart disease [1]. Unexpectedly high incidence of thrombosis have been reported [2, 3], and severity of COVID-19 disease is associated with elevated inflammatory markers and markers of coagulation such as d-Dimer, fibrinogen and von Willebrand factor [1, 4]. Moreover, COVID-19 autopsies have shown evidence of widespread microthrombosis in the lungs and other organs [5].

Circulating platelets play a central role in haemostasis and thrombosis, and platelets significantly contribute to immune responses during viral infection in a process termed “immunothrombosis” [6]. COVID-19 patients have higher levels of P-selectin expression in resting and activated platelets, elevated circulating platelet-leukocyte aggregates, increased aggregation, and thromboxane generation [7, 8]. Platelet hyperreactivity may contribute to immunothrombosis often seen in patients with COVID-19 [9]. In addition, mild thrombocytopenia is observed in COVID-19 patients, and a progressive decline of platelet counts was significantly associated with increased mortality [10]. Moreover, pulmonary megakaryocytes are increased in COVID-19 patients with acute lung injury [11]. Since the lung is considered an active site of megakaryopoiesis, a prothrombotic status leading to platelet activation, aggregation and consumption may trigger a compensatory pulmonary response [11].

Platelet activation markers are useful tools in evaluating risk factors of thrombosis in a variety of clinical conditions such as acute myocardial infarction, atherosclerosis, type 2 diabetes mellitus and other inflammatory diseases [12]. While there are many methods used to test platelet activation for research purposes, most of the existing techniques are expensive, require trained personnel and take time to perform, limiting their use in clinical practice [12].

Circulation of larger, younger platelets reflect platelets activity and seem to be useful predictors and prognostic biomarker of thrombotic events [13, 14]. Platelet size can be assessed during a routine clinical blood test using automated haematology analysers. There are several methods on automated analysers for measuring platelet size and count, including aperture impedance, optical scattering, and fluorescence [15]. Platelet morphological parameters include mean platelet volume (MPV), platelet distribution width (PDW) and platelet-large cell ratio (P-LCR) (Table 1). The aim of this rapid evidence review is to assess the potential association of increased MPV, PDW and P-LCR with disease severity and mortality in patients with COVID-19.

**Table 1:**
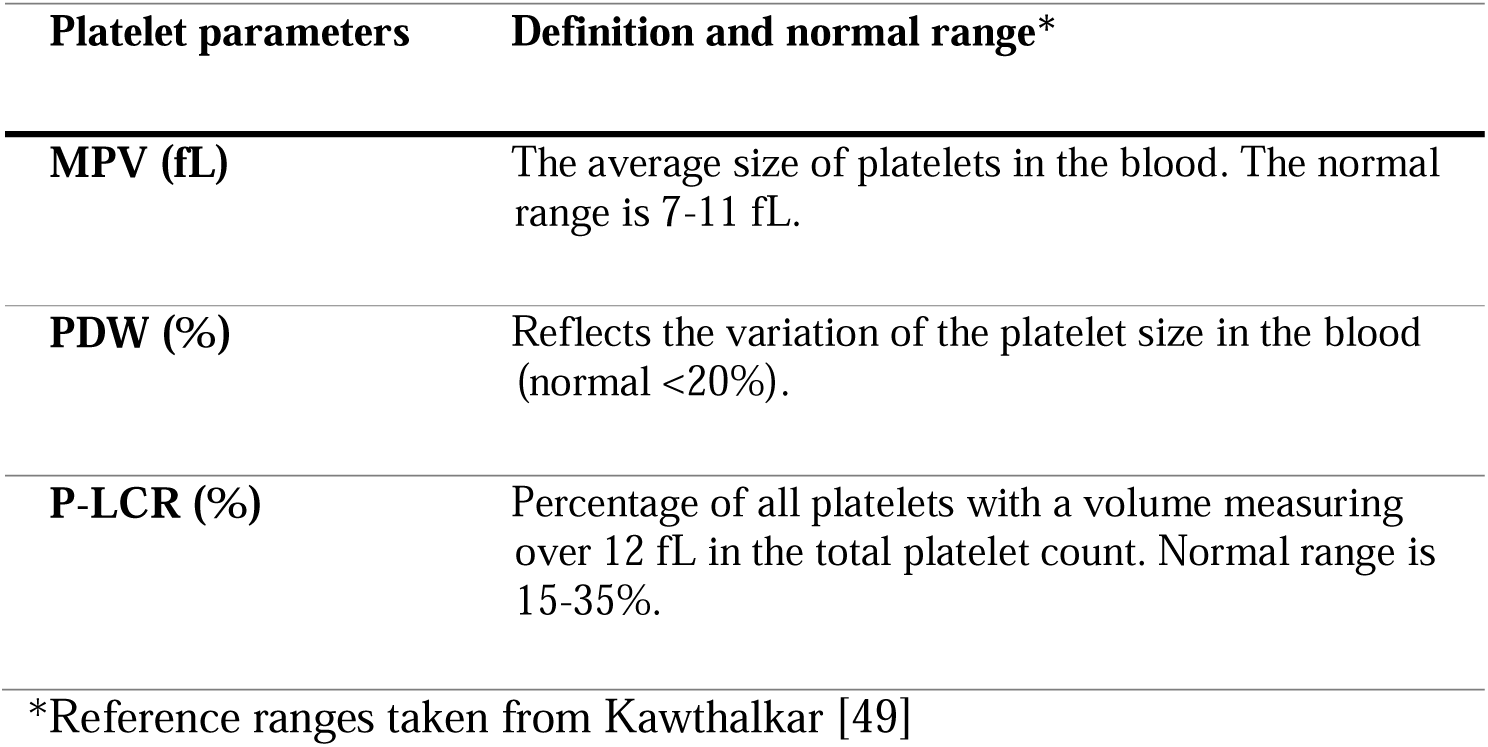
Description of platelet parameters reflecting platelet size

| **Platelet parameters** | **Definition and normal range\*** |
| --- | --- |
| **MPV (fL)** | The average size of platelets in the blood. The normal range is 7-11 fL. |
| **PDW (%)** | Reflects the variation of the platelet size in the blood (normal <20%). |
| **P-LCR (%)** | Percentage of all platelets with a volume measuring over 12 fL in the total platelet count. Normal range is 15-35%. |

\*Reference ranges taken from Kawthalkar [49]

## METHODS

### Search Strategy

A review protocol was published on PROSPERO (ID: CRD42021242848). The review is reported in line with the Preferred Reporting Items for Systematic Reviews and Meta-Analyses (PRISMA) guidelines. We carried out a systematic search of the literature from Medline, Embase, PubMed, Web of Science, the Cochrane Central Register of Controlled Trials (CENTRAL) for all literature published up to 27th March 2021. Searches were limited to English language. Relevant studies were identified for all reported studies of associations between COVID-19 and platelet indices reflecting platelet size using the terms: “covid” OR “coronavirus” OR “ncov” OR “sars” OR “sars-cov” AND “mean platelet volume” OR “platelet distribution width” OR “platelet large cell ratio”. A search of the preprint databases, MedRixv and BioRixv, was conducted for all literature published from 1st January 2020 to 7th April 2021’ using phrase terms for “mean platelet volume”, “platelet distribution width” and “platelet large cell ratio”. The China Knowledge Resource Integrated (CNKI) database was searched for literature up to 25^th^ April 2021, using the search terms “血小板(platelet)” AND “COVID-19”. Hand searching was also performed in the reference lists of relevant articles to identify additional eligible studies. See Supplementary material 1 for details of the search strategy.

### Inclusion/exclusion criteria

Studies were included in this review if they met the criteria as follows:

Inclusion criteria: 1) Adult patients with laboratory-confirmed COVID-19; 2) biomarker reflecting platelet size (i.e., MPV, PDW and/or P-LCR); 3) investigation of an association between a biomarker reflecting platelet size and disease severity and/or mortality in COVID-19; 4) original (experimental) research including randomised controlled trials, case-control studies, cohort studies, cross-sectional, case reports and series of cases.; 5) articles in English or Chinese language.

Exclusion criteria: 1) Under 18-year-olds 2) Reviews, meta-analyses, conference abstracts, editorials, guidelines, commentaries, protocols 3) animal-based experiments; 4) *in vitro* studies; 5) unrelated studies; 6) studies focused on specific patient populations e.g. diabetic or cancer patients.

### Study selection

All records identified by the database search were screened by title and abstract. A random sample of 20% of the title/abstracts were screened from the English literature and discussed between two authors (SD and HW), and the remaining abstracts were screened by SD. Chinese literature was translated into English by the Chinese speaking reviewer, HW, and a random sample of 20% was screened by SD and HW. The remaining title/abstracts from the Chinese literature were screened by HW. Studies considered relevant were evaluated in full text according to the prespecified inclusion and exclusion criteria. The full text article from the Chinese literature was translated into English by HW. A random sample of 20% of the full text articles were screened for the English language literature and discussed between two authors (SD and HW), and the remaining full text articles were screened by SD.

### Data extraction

One reviewer (SD) extracted data from each study and compiled summary tables. A second reviewer (HW) randomly selected about 50% of the data extraction to check the accuracy. Any discrepancies identified were discussed and resolved between SD and HW and reflected in the remaining 50%. A second reviewer (HW) verified 50% of the data extraction. For all included studies, the following data was extracted: lead author, publication year, country, study design, study population (including age and % females), sample number, severity definition, day/time of blood test, subject exclusion if anti-platelet medication taken ≤10 days prior to test (yes or no), follow-up time (cohort studies only), primary outcomes for the meta-analysis.

The primary outcomes in our meta-analysis were the correlation of MPV, PDW or P-LCR parameters on severity or mortality. We included the platelet count (PLT) for qualitative analysis. We selected for the severity definitions: mild, moderate, severe and critical. Endpoint measures using mean ± SD, median [min-max] or median [interquartile range (IQR)] were extracted for platelet indices. For data consistency, studies reporting measures as ‘changes from baseline’ were excluded from the meta-analysis. If results were expressed as median [min-max] or median [IQR], the mean ± SD of MPV was acquired by contacting the corresponding authors or by converting using the formula from Shi et al [16]. If the median [min-max] or median [IQR] data were skewed, the studies were excluded from the meta-analysis to avoid misleading or unreliable conclusions for the transformed mean and SD. We contacted the authors of 13 studies to request data that was not directly available from the literature and five authors provided sufficient data for meta-analysis. When patient data was available for more than one day, we took the data from blood tests taken within 24 hours of hospital admission.

### Quality assessment

All included articles were quality assessed using the National Institutes of Health’s Quality Assessment Tools for Observational Cohort and Cross-Sectional Studies [17]. The questionnaire items were discussed by two reviewers (SD and HW) prior to conducting the quality assessment. It was decided that the study quality would be judged based on its association with this study, rather than as an independent study. For example, if the study predominantly used *in vitro* methods for analysing platelet activation, we only judged the study based on its methods for collecting and analysing clinical laboratory data.

The quality assessment was independently conducted by one reviewer (SD). A second reviewer (HW) randomly selected about 50% of the data extraction to check the accuracy. Any discrepancies identified were discussed and resolved between SD and HW and reflected in the remaining 50%. Each study was rated as poor, fair or good based on the details that were reported and consideration of the concepts for minimizing bias.

### Data analysis

The mean values of MPV, PDW and P-LCR between the non-severe vs severe and survivor vs. non-survivor groups were estimated and pooled using the standardized mean difference (SMD). We employed a random-effects and inverse-variance weighting using Review Manager (RevMan v5.4.1 2020). Heterogeneity between the studies was assessed using the *I*^2^ statistic. We performed a sensitivity analysis based on the day the blood test was performed (specifically blood tests taken at hospital admission i.e. day 0) and clinical outcomes, if appropriate. A subgroup analysis was performed based on the quality assessment rating of poor or fair/good and studies reporting clear or unclear outcome measures. Although there is no universally accepted optimal minimum number of studies that are required for a subgroup analysis, we performed the analysis only if ≥6 studies could be applied to each subgroup based on Fu et al [18]. Publication bias was examined using a funnel plot if there were ≥10 studies in the meta-analysis [19]. All the statistical heterogeneity was assessed using RevMan v5.4.1 (2020). For unreported *p* values, an appropriate t-test was performed using Graphpad v9.1.1 (2021) if raw data was available. A *p* value < 0.05 was considered statistically significant.

To explain the obtained Cohen’s d in a common language, the common language effect size (CLES) was calculated for studies that tested the platelet size at hospital admission. For this study, the CLES represents the probability that a person selected at random from the severe/non-survivor group will have a higher platelet size (e.g. MPV, PDW or P-LCR) than a person selected at random from the non-severe/survived group (%): CLES = Φ (Cohen’s d / √ 2), where Φ is the cumulative distribution function of the standard normal distribution.

## RESULTS

### Study characteristics

We identified a total of 80 records from the OVID (Medline and Embase) search, 45 records from the Web of Science database, 49 records from PubMed, 3 records from CENTRAL, 133 records from the MedRxiv database, 38 from CNKI and 3 records from references searches (Figure 1). Of these, 120 were duplicates. Two-hundred and thirty-two records were title and abstract screened, and 46 were taken to full-text review. Thirty-five studies were included for the data collection stage, and 13 were excluded due to insufficient outcome data. Twenty-two studies (11,906 patients) were included in the meta-analysis (Figure 1).

**Figure 1:**
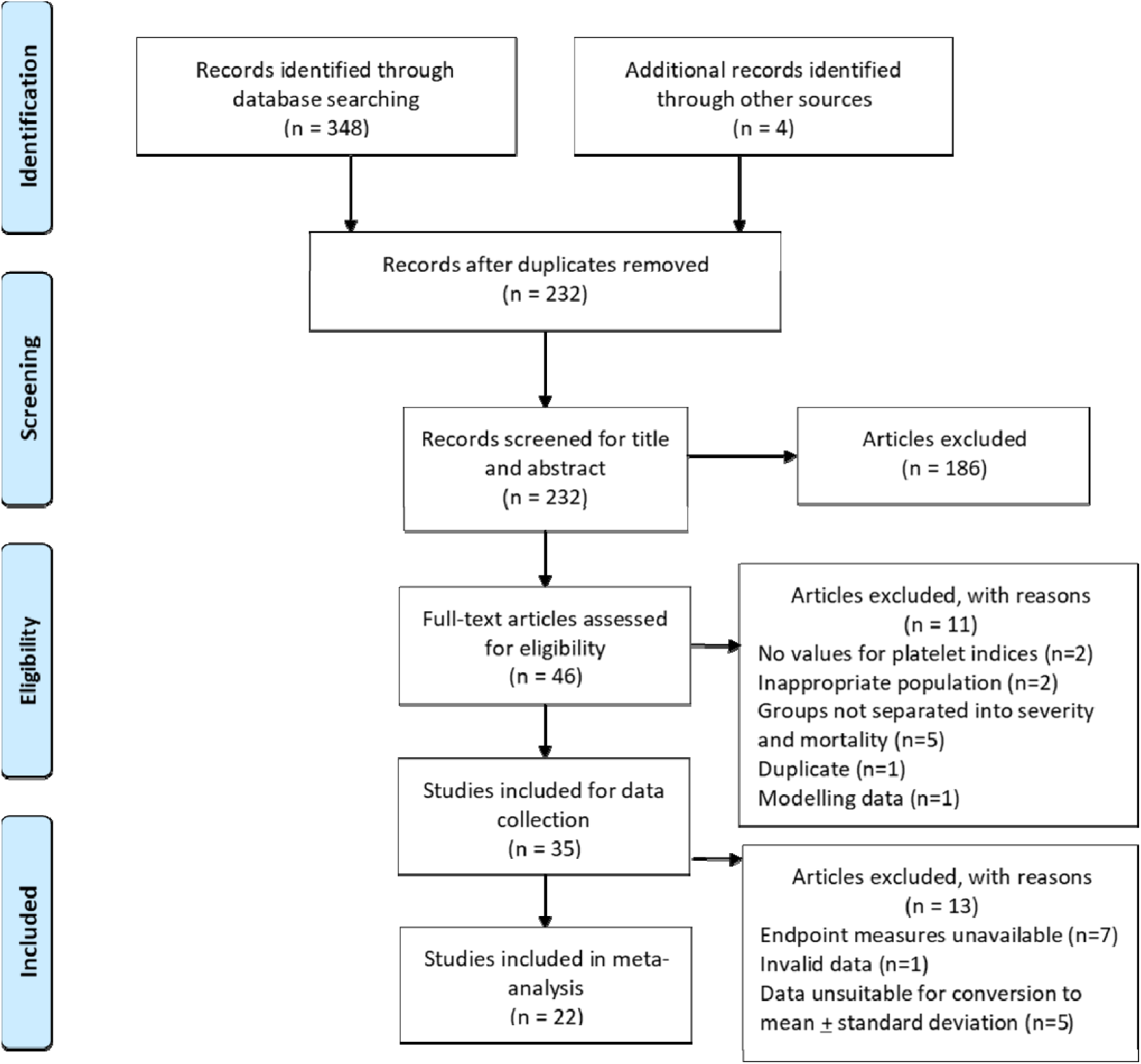
PRISMA diagram of the search strategy.

The baseline characteristics of the included studies are presented in Table 2. Nineteen studies were retrospective, and three were prospective, observational studies. The largest number of studies were from China (n=9), and most were single-centre studies (n=16). Wu et al [20] was an international multi-centre study, but we extracted data for patients recruited from one centre. Five studies were identified from preprint databases [20-24]. The disease outcomes for 16 studies were severity of COVID-19, and seven studies assessed mortality. One study provided a comparison between groups for disease severity and mortality combined [25].

**Table 2:**
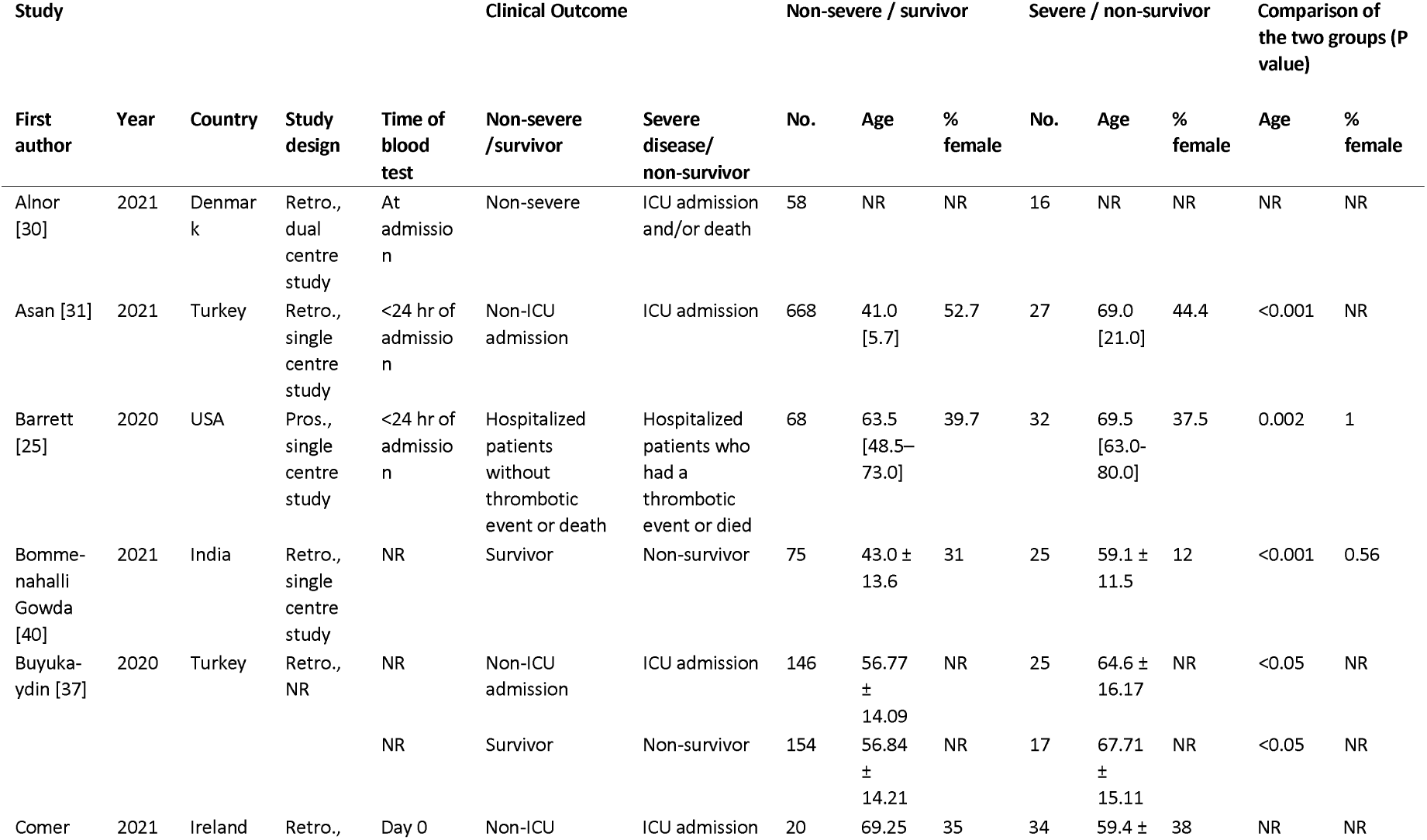
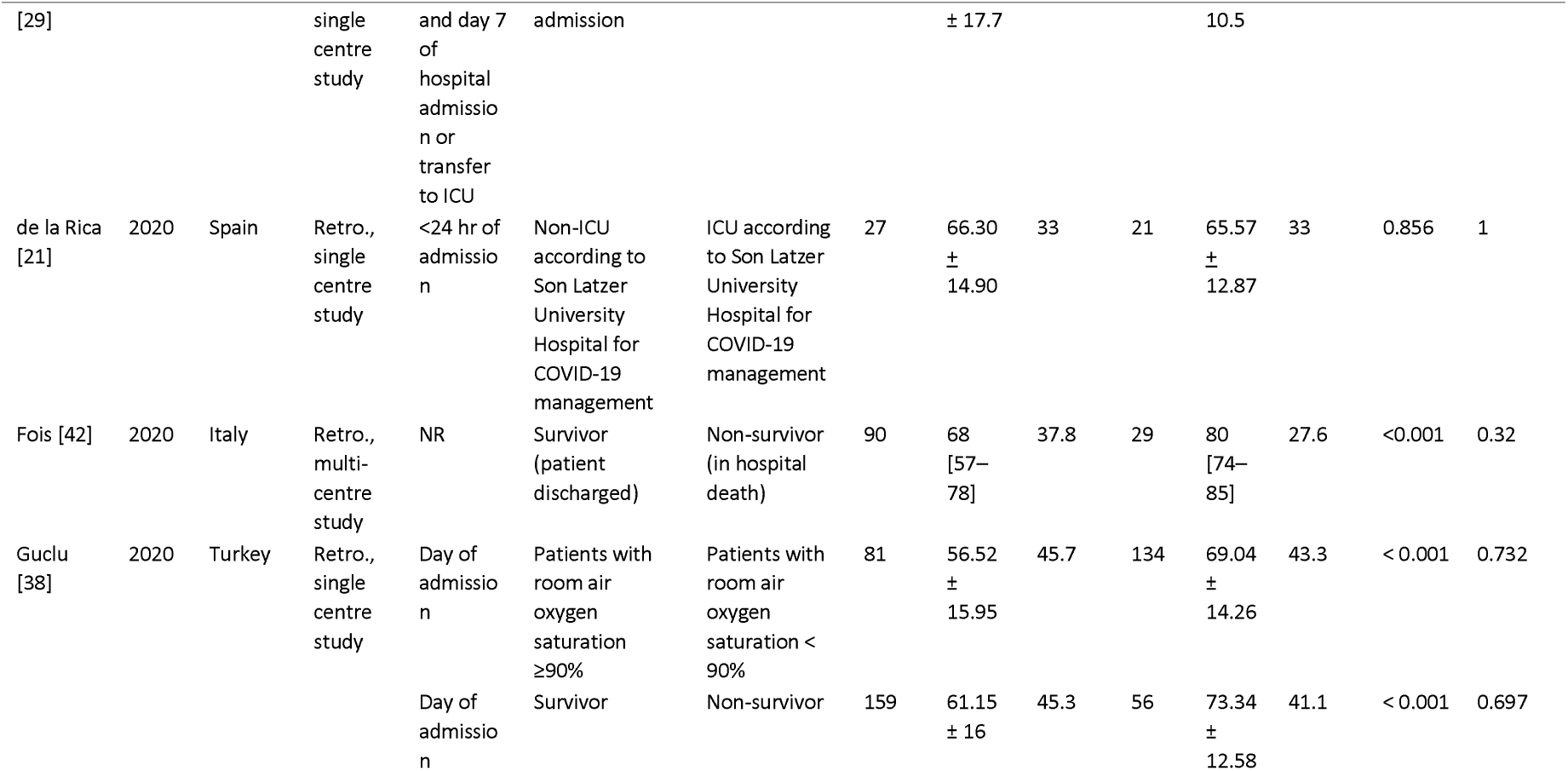
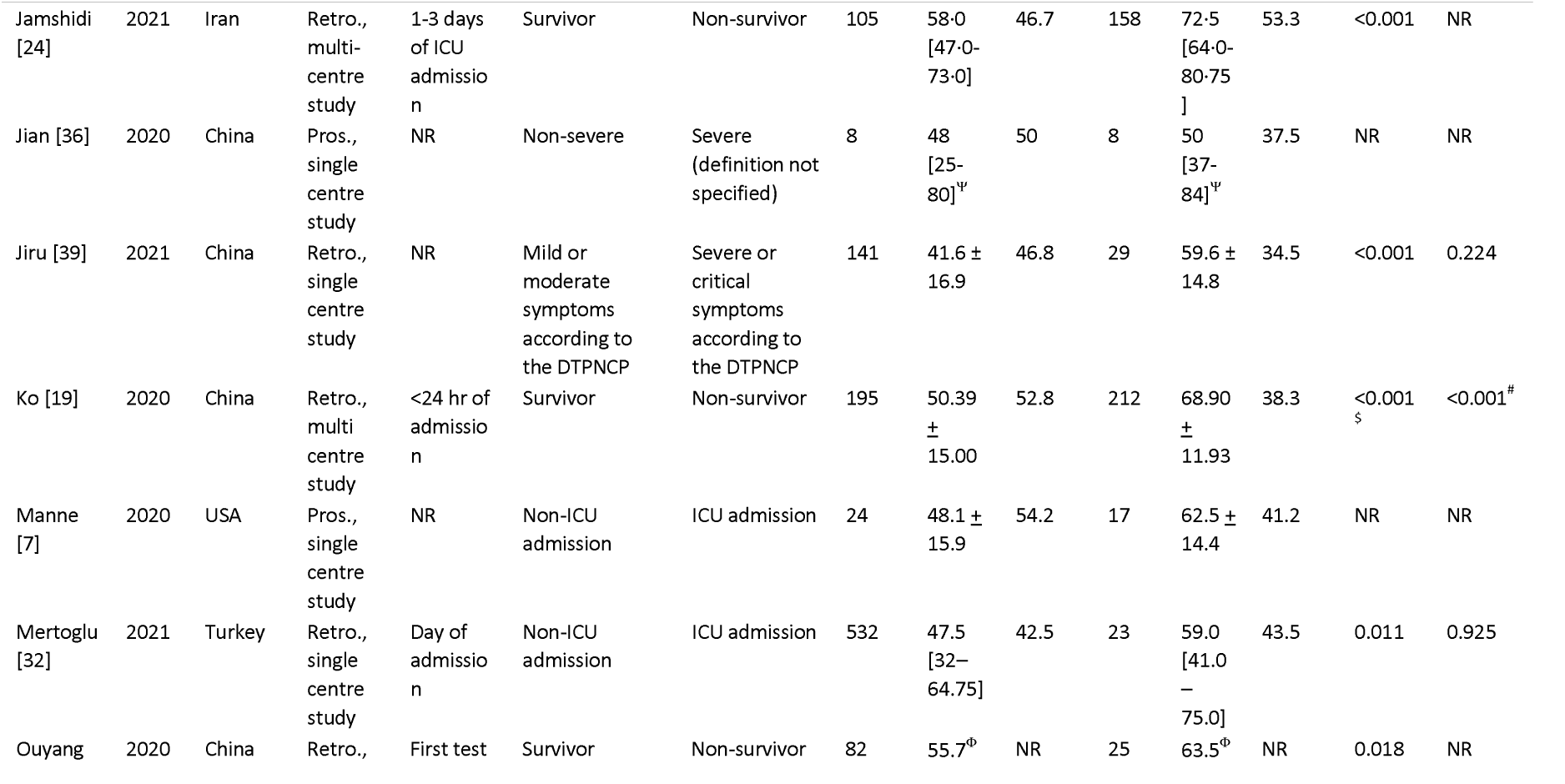
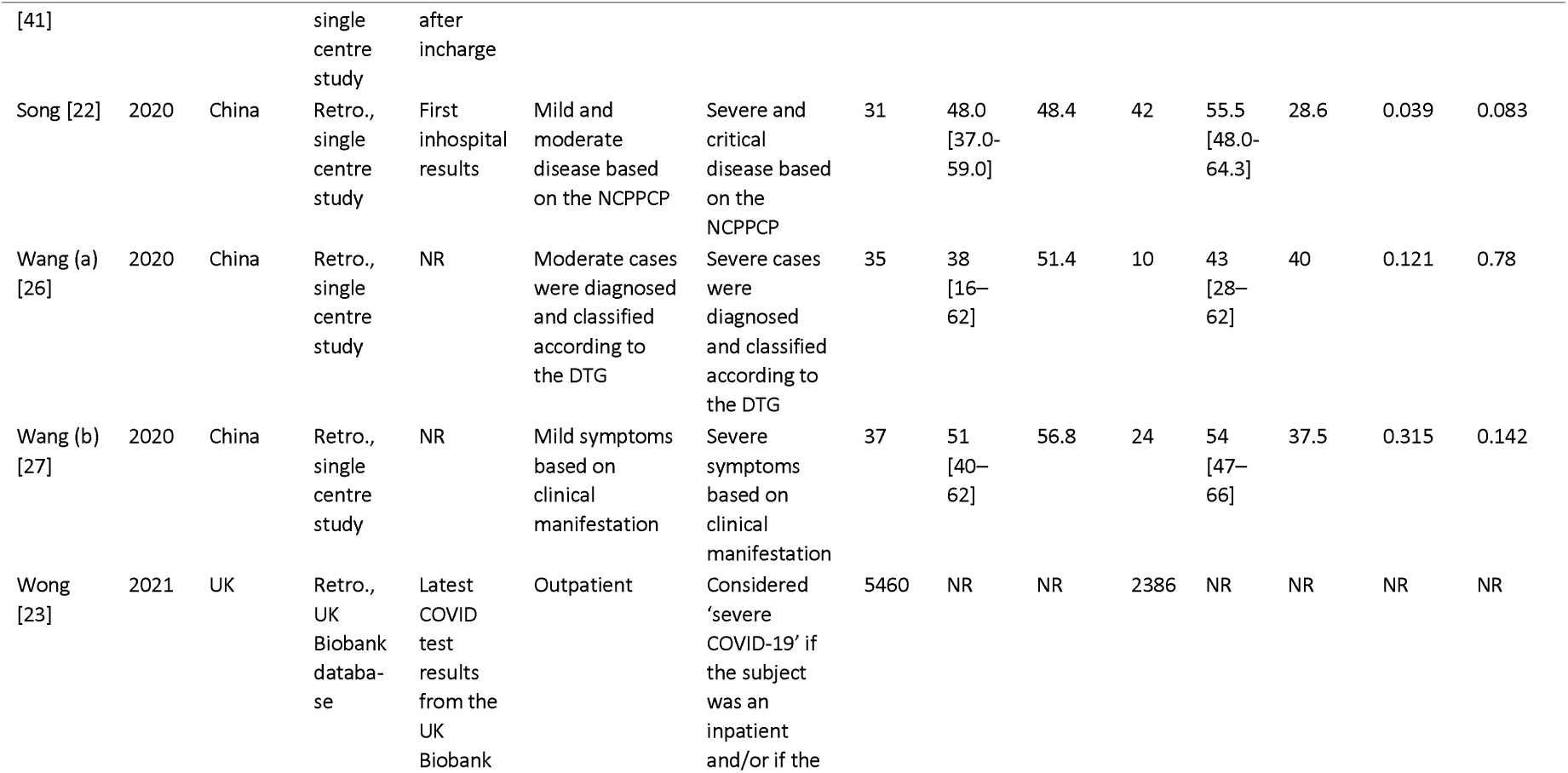
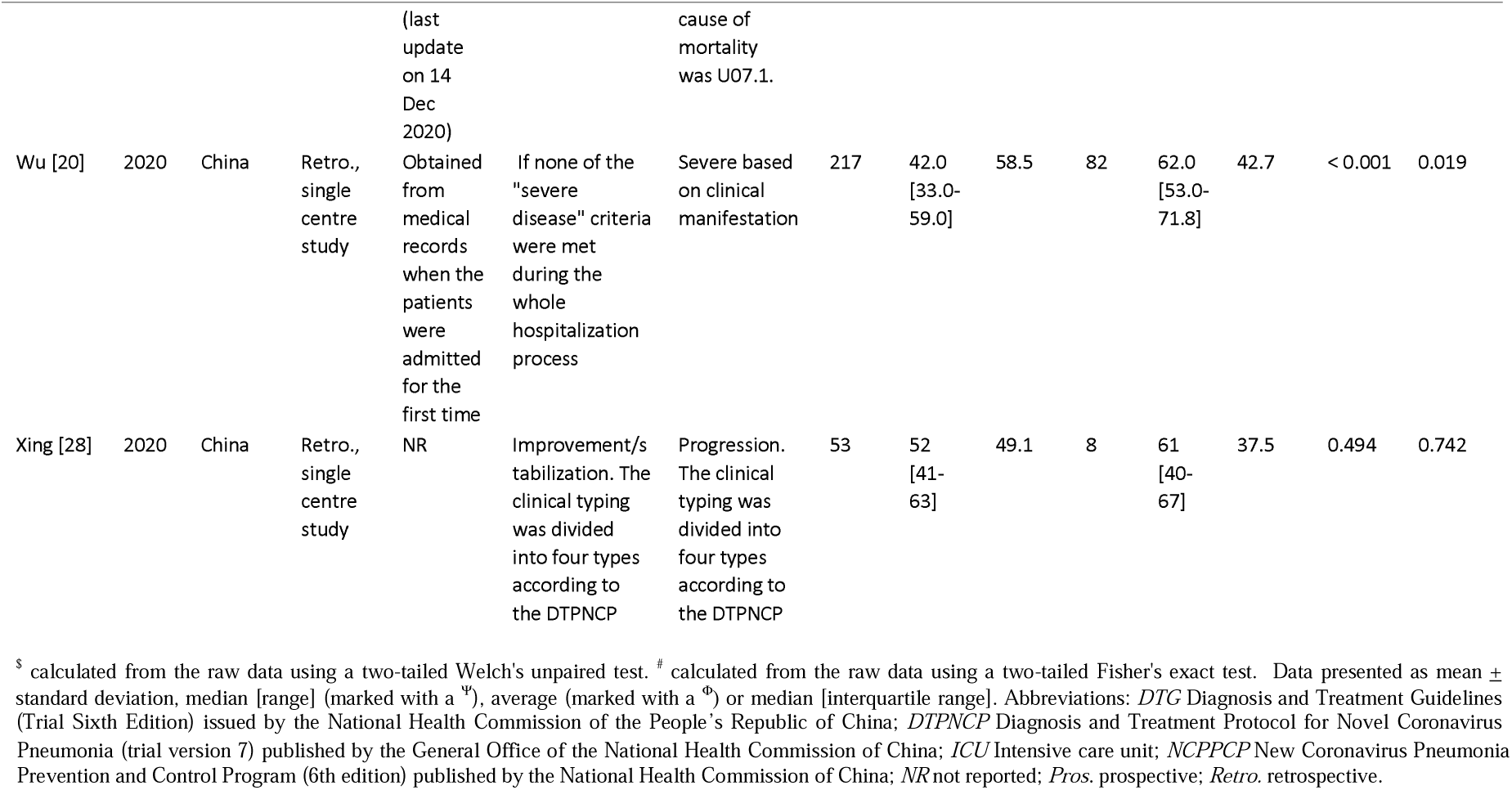
Study characteristics

| Study |  |  |  |  | Clinical Outcome |  | Non-severe / survivor |  |  | Severe / non-survivor |  |  | Comparison of the two groups (P value) |  |
| --- | --- | --- | --- | --- | --- | --- | --- | --- | --- | --- | --- | --- | --- | --- |
| First author | Year | Country | Study design | Time of blood test | Non-severe /survivor | Severe disease/ non-survivor | No. | Age | % female | No. | Age | % female | Age | % female |
| Alnor [30] | 2021 | Denmark | Retro., dual centre study | At admission | Non-severe | ICU admission and/or death | 58 | NR | NR | 16 | NR | NR | NR | NR |
| Asan [31] | 2021 | Turkey | Retro., single centre study | <24 hr of admission | Non-ICU admission | ICU admission | 668 | 41.0 [5.7] | 52.7 | 27 | 69.0 [21.0] | 44.4 | <0.001 | NR |
| Barrett [25] | 2020 | USA | Pros., single centre study | <24 hr of admission | Hospitalized patients without thrombotic event or death | Hospitalized patients who had a thrombotic event or died | 68 | 63.5 [48.5–73.0] | 39.7 | 32 | 69.5 [63.0–80.0] | 37.5 | 0.002 | 1 |
| Bommenahalli Gowda [40] | 2021 | India | Retro., single centre study | NR | Survivor | Non-survivor | 75 | 43.0 ± 13.6 | 31 | 25 | 59.1 ± 11.5 | 12 | <0.001 | 0.56 |
| Buyukaydin [37] | 2020 | Turkey | Retro., NR | NR | Non-ICU admission | ICU admission | 146 | 56.77 ± 14.09 | NR | 25 | 64.6 ± 16.17 | NR | <0.05 | NR |
|  |  |  |  | NR | Survivor | Non-survivor | 154 | 56.84 ± 14.21 | NR | 17 | 67.71 ± 15.11 | NR | <0.05 | NR |
| Comer | 2021 | Ireland | Retro., | Day 0 | Non-ICU | ICU admission | 20 | 69.25 | 35 | 34 | 59.4 ± | 38 | NR | NR |

| Study |  |  |  | Clinical Outcome |  |  | Non-severe / survivor |  |  | Severe / non-survivor |  |  | Comparison of the two groups (P value) |  |
| --- | --- | --- | --- | --- | --- | --- | --- | --- | --- | --- | --- | --- | --- | --- |
| First author | Year | Country | Study design | Time of blood test | Non-severe /survivor | Severe disease/ non-survivor | No. | Age | % female | No. | Age | % female | Age | % female |
| [29] |  |  | single centre study | and day 7 of hospital admission or transfer to ICU | admission |  |  | ± 17.7 |  |  | 10.5 |  |  |  |
| de la Rica [21] | 2020 | Spain | Retro., single centre study | <24 hr of admission | Non-ICU according to Son Latzer University Hospital for COVID-19 management | ICU according to Son Latzer University Hospital for COVID-19 management | 27 | 66.30 ± 14.90 | 33 | 21 | 65.57 ± 12.87 | 33 | 0.856 | 1 |
| Fois [42] | 2020 | Italy | Retro., multi-centre study | NR | Survivor (patient discharged) | Non-survivor (in hospital death) | 90 | 68 [57–78] | 37.8 | 29 | 80 [74–85] | 27.6 | <0.001 | 0.32 |
| Guclu [38] | 2020 | Turkey | Retro., single centre study | Day of admission | Patients with room air oxygen saturation ≥90% | Patients with room air oxygen saturation < 90% | 81 | 56.52 ± 15.95 | 45.7 | 134 | 69.04 ± 14.26 | 43.3 | < 0.001 | 0.732 |
|  |  |  |  | Day of admission | Survivor | Non-survivor | 159 | 61.15 ± 16 | 45.3 | 56 | 73.34 ± 12.58 | 41.1 | < 0.001 | 0.697 |
| First author | Year | Country | Study design | Time of blood test | Non-severe /survivor | Severe disease/ non-survivor | No. | Age | % female | No. | Age | % female | Age | % female |
| Jamshidi [24] | 2021 | Iran | Retro., multi-centre study | 1-3 days of ICU admission | Survivor | Non-survivor | 105 | 58.0 [47.0-73.0] | 46.7 | 158 | 72.5 [64.0-80.75] | 53.3 | <0.001 | NR |
| Jian [36] | 2020 | China | Pros., single centre study | NR | Non-severe | Severe (definition not specified) | 8 | 48 [25-80]<sup>ψ</sup> | 50 | 8 | 50 [37-84]<sup>ψ</sup> | 37.5 | NR | NR |
| Jiru [39] | 2021 | China | Retro., single centre study | NR | Mild or moderate symptoms according to the DTPNCP | Severe or critical symptoms according to the DTPNCP | 141 | 41.6 ± 16.9 | 46.8 | 29 | 59.6 ± 14.8 | 34.5 | <0.001 | 0.224 |
| Ko [19] | 2020 | China | Retro., multi centre study | <24 hr of admission | Survivor | Non-survivor | 195 | 50.39 ± 15.00 | 52.8 | 212 | 68.90 ± 11.93 | 38.3 | <0.001<sup>§</sup> | <0.001<sup>#</sup> |
| Manne [7] | 2020 | USA | Pros., single centre study | NR | Non-ICU admission | ICU admission | 24 | 48.1 ± 15.9 | 54.2 | 17 | 62.5 ± 14.4 | 41.2 | NR | NR |
| Mertoglu [32] | 2021 | Turkey | Retro., single centre study | Day of admission | Non-ICU admission | ICU admission | 532 | 47.5 [32–64.75] | 42.5 | 23 | 59.0 [41.0–75.0] | 43.5 | 0.011 | 0.925 |
| Ouyang | 2020 | China | Retro., | First test | Survivor | Non-survivor | 82 | 55.7<sup>Φ</sup> | NR | 25 | 63.5<sup>Φ</sup> | NR | 0.018 | NR |
| First author | Year | Country | Study design | Time of blood test | Non-severe /survivor | Severe disease/ non-survivor | No. | Age | % female | No. | Age | % female | Age | % female |
| [41] |  |  | single centre study | after incharge |  |  |  |  |  |  |  |  |  |  |
| Song [22] | 2020 | China | Retro., single centre study | First in-hospital results | Mild and moderate disease based on the NCPPCP | Severe and critical disease based on the NCPPCP | 31 | 48.0 [37.0-59.0] | 48.4 | 42 | 55.5 [48.0-64.3] | 28.6 | 0.039 | 0.083 |
| Wang (a) [26] | 2020 | China | Retro., single centre study | NR | Moderate cases were diagnosed and classified according to the DTG | Severe cases were diagnosed and classified according to the DTG | 35 | 38 [16-62] | 51.4 | 10 | 43 [28-62] | 40 | 0.121 | 0.78 |
| Wang (b) [27] | 2020 | China | Retro., single centre study | NR | Mild symptoms based on clinical manifestation | Severe symptoms based on clinical manifestation | 37 | 51 [40-62] | 56.8 | 24 | 54 [47-66] | 37.5 | 0.315 | 0.142 |
| Wong [23] | 2021 | UK | Retro., UK Biobank database | Latest COVID test results from the UK Biobank | Outpatient | Considered 'severe COVID-19' if the subject was an inpatient and/or if the | 5460 | NR | NR | 2386 | NR | NR | NR | NR |

| Study |  |  |  | Clinical Outcome |  |  | Non-severe / survivor |  |  | Severe / non-survivor |  |  | Comparison of the two groups (P value) |  |
| --- | --- | --- | --- | --- | --- | --- | --- | --- | --- | --- | --- | --- | --- | --- |
| First author | Year | Country | Study design | Time of blood test | Non-severe /survivor | Severe disease/ non-survivor | No. | Age | % female | No. | Age | % female | Age | % female |
|  |  |  |  | (last update on 14 Dec 2020) |  | cause of mortality was U07.1. |  |  |  |  |  |  |  |  |
| Wu [20] | 2020 | China | Retro., single centre study | Obtained from medical records when the patients were admitted for the first time | If none of the "severe disease" criteria were met during the whole hospitalization process | Severe based on clinical manifestation | 217 | 42.0 [33.0-59.0] | 58.5 | 82 | 62.0 [53.0-71.8] | 42.7 | < 0.001 | 0.019 |
| Xing [28] | 2020 | China | Retro., single centre study | NR | Improvement/s tabilization. The clinical typing was divided into four types according to the DTPNCP | Progression. The clinical typing was divided into four types according to the DTPNCP | 53 | 52 [41-63] | 49.1 | 8 | 61 [40-67] | 37.5 | 0.494 | 0.742 |

<sup>§</sup> calculated from the raw data using a two-tailed Welch's unpaired test. <sup>#</sup> calculated from the raw data using a two-tailed Fisher's exact test. Data presented as mean ± standard deviation, median [range] (marked with a <sup>ψ</sup>), average (marked with a <sup>Φ</sup>) or median [interquartile range]. Abbreviations: *DTG* Diagnosis and Treatment Guidelines (Trial Sixth Edition) issued by the National Health Commission of the People's Republic of China; *DTPNCP* Diagnosis and Treatment Protocol for Novel Coronavirus
Pneumonia (trial version 7) published by the General Office of the National Health Commission of China; *ICU* Intensive care unit; *NCPPCP* New Coronavirus Pneumonia Prevention and Control Program (6th edition) published by the National Health Commission of China; *NR* not reported; *Pros.* prospective; *Retro.* retrospective.

Most studies classified disease severity as patients who were admitted to an intensive care unit (ICU) or presented with at least one of the clinical manifestations listed in national guidelines for severe or critical diagnosis of COVID-19. Only four studies recruited subjects with similar ages between groups (p >0.05) [21, 26-28], and two studies had significantly different male:female ratios (p <0.05) [19, 20]. One study excluded patients on antiplatelet drugs >10 days prior to the blood test [29] and another study adjusted for antiplatelet therapy in the data analysis [25]. Values for platelet indices reported by each study are listed in Table 3. Although there was a trend toward lower PLT in the severe and non-survivor groups for most studies, it was only significantly lower in three studies of severe patients [20, 29, 30] and two studies of non-survivors [19, 24]. Only one study reported a mean PLT for the severe COVID-19 patients that was within the mild thrombocytopenia range (<150 ×10^9^) [20].

**Table 3:** Values of platelet indices in the COVID-19 patient severe and non-severe groups

| First author | Year | Platelet indices | Non-severe patients | Severe patients | Comparison of the two groups (P value) |
| --- | --- | --- | --- | --- | --- |
| Alnor [30] | 2021 | PLT | 188 [161–271] | 174 [127–203] | 0.037 |
|  |  | MPV | 10.5 [9.6–10.9] | 10.9 [10.1–11.8] | 0.126 |
|  |  | PDW | 12.1 [10.5–13.2] | 13.0 [11.6–14.5] | 0.097 |
|  |  | P-LCR | 28.5 [22.9–32.7] | 31.1 [25.9–39.4] | 0.093 |
| Asan [31] | 2021 | PLT | 221 ± 60 | 198 ± 61 | 0.081 |
|  |  | MPV | 9.9 ± 4.2 | 10.1 ± 1.6 | 0.241 |
|  |  | MPV | 10.55 [10.1–11.2] | 11.00 [10.5–11.9] | 0.022 |
|  |  | PDW | 16.37 ± 2.59 | 17.58 ± 2.84 | 0.05 |
| Buyukaydin [37] | 2020 | PLT | 219.12 ± 79.25 | 249.56 ± 88.26 | 0.087 |
|  |  | MPV | 7.82 ± 1.39 | 7.77 ± 1.12 | 0.946 |
| Comer [29] | 2021 | PLT | 322 [122–513]<sup>o</sup> | 235 [36–550]<sup>o</sup> | 0.014 |
|  |  | MPV | 9.6 [8.5–11.7]<sup>o</sup> | 10.8 [9–13.2]<sup>o</sup> | 0.0014 |
| de la Rica [21] | 2020 | PLT | 228.02 ± 108.82 | 209.56 ± 77.97 | 0.678 |
|  |  | MPV | 7.92 ± 1.25 | 8.18 ± 1.19 | 0.596 |
|  |  | PDW | 16.81 ± 1.24 | 16.99 ± 0.97 | 0.333 |
| Guclu [38] | 2020 | PLT | 187.4 ± 59.82 | 208.63 ± 135.72 | 0.573 |
|  |  | MPV | 9.18 ± 1.24 | 9.61 ± 1.76 | 0.129 |
|  |  | PDW | 17.37 ± 2.32 | 17.72 ± 2.52 | 0.142 |
| Jian [36] | 2020 | PLT | 219.25 + 80.35 | 152 + 42.85 | <0.05 |
|  |  | MPV | 10.93 + 1.79 | 11.64 + 1.52 | NS |
| Jiru [39] | 2021 | PLT | 184.3 ± 58.5 | 173.9 ± 56.5 | 0.173 |
|  |  | MPV | 11.0 ± 1.2 | 10.9 ± 1.2 | 0.182 |
|  |  | PDW | 14.8 ± 3.2 | 15.3 ± 2.1 | 0.84 |
| Manne [7] | 2020 | PLT | 252.5 ± 112.1 | 238.7 ± 59.9 | NR |
|  |  | MPV | 9.84 ± 0.71 | 10.34 ± 0.77 | NR |
| Mertoglu [32] | 2021 | PLT | 233.0 [193.0-278.0] | 219.0 [176.0-312.0] | 0.872 |
|  |  | MPV | 10.1 [9.5-10.7] | 10.8 [9.6-11.7] | 0.052 |
|  |  | P-LCR | 25.60 [21.0-30.61] | 32.70 [22.8-38.0] | 0.033 |
| Song [22] | 2020 | PLT | 178.0 [127.0-239.0] | 189.0 [154.0-231.0] | 0.321 |
|  |  | PDW | 12.8 [10.7-13.9] | 12.3 [11.0-13.5] | 0.643 |
| Wang (a) [26] | 2020 | PLT | 222.7 ± 73.01 | 210.6 ± 60.43 | 0.403 |
|  |  | MPV | 9.74 ± 1.22 | 10.17 ± 0.98 | 0.075 |
|  |  | PDW | 16.23 ± 0.31 | 16.36 ± 0.30 | 0.04 |
|  |  | P-LCR | 24.55 ± 8.11 | 27.20 ± 6.54 | 0.097 |
| Wang (b) [27] | 2020 | PLT | 217.0 [18.3-252.0] | 163.5 [133.25-248.25] | 0.066 |
|  |  | MPV | 10.1 [9-10.8] | 10.4 [9.65-11.18] | 0.243 |
| Wong [23] | 2021 | PLT | 253.68 [59.10] | 250.91 [63.08] | NR |
|  |  | MPV | 9.36 [1.06] | 9.33 [1.11] | NR |
| Wu [20] | 2020 | PLT | 174.0 [140.8-214.5] | 149.0 [116.5-188.8] | <0.001 |
|  |  | MPV | 9.9 [9.2-10.0] | 10.0 [9.2-10.0] | 0.146 |
|  |  | P-LCR | 24.3 [19.9-30.0] | 25.4 [19.7-31.0] | 0.192 |
| Xing [28] | 2020 | PLT | 212.0 [163.0-252.0] | 174.0 [135.5-239.0] | 0.207 |
|  |  | MPV | 10.10 [9.00-10.95] | 10.10 [9.70-11.60] | 0.494 |

Data presented as mean ± standard deviation, median [min-max range] (marked with a ω) or median [interquartile range]. Units for platelet indices are PLT: 10<sup>9</sup>/L; MPV: fL; PDW: %; P=LCR: %. Abbreviations: MPV mean platelet volume; NS not significant; P-LCR platelet-large cell ratio; PLT platelet count; PDW platelet distribution width

**Table 4:**
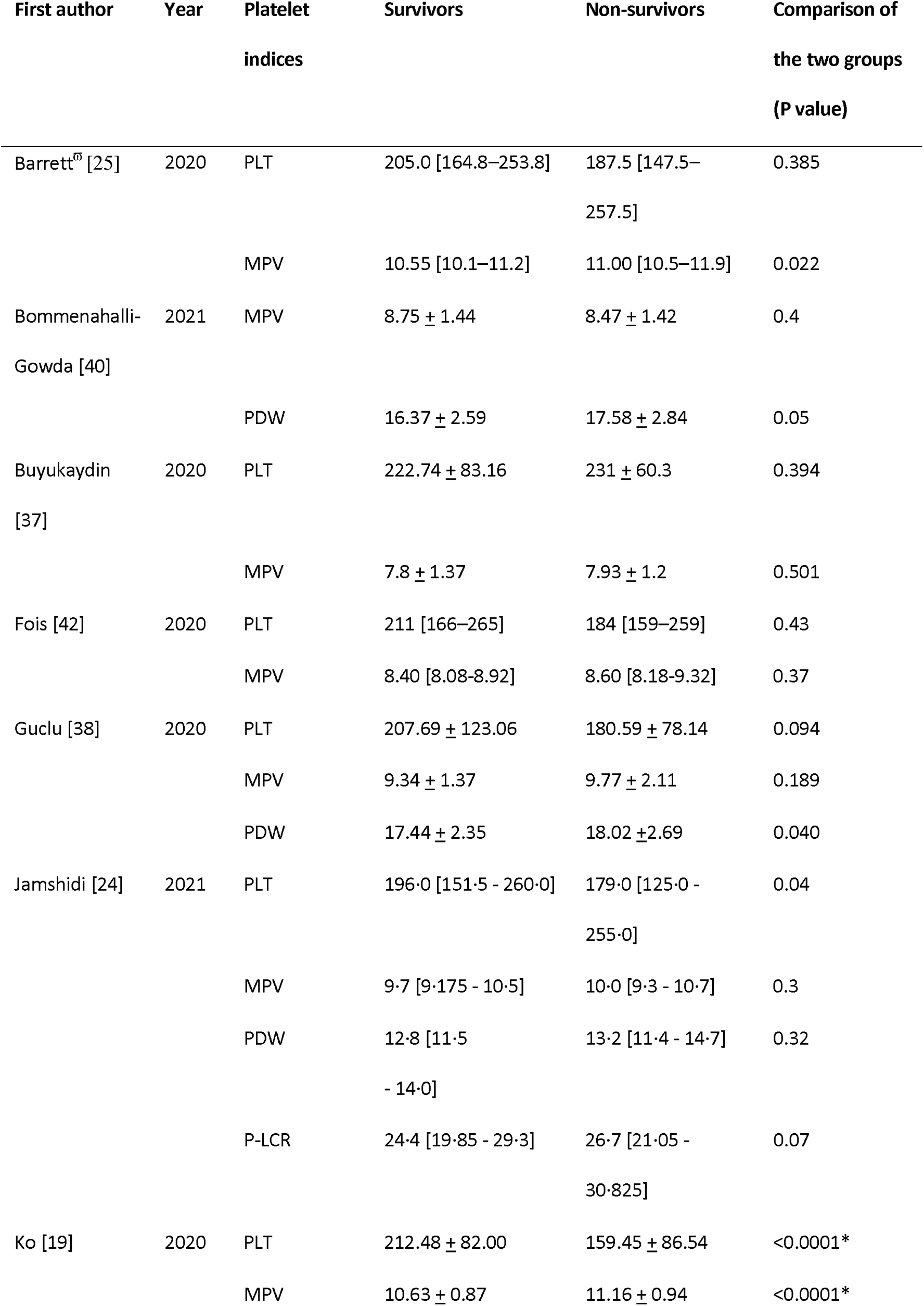
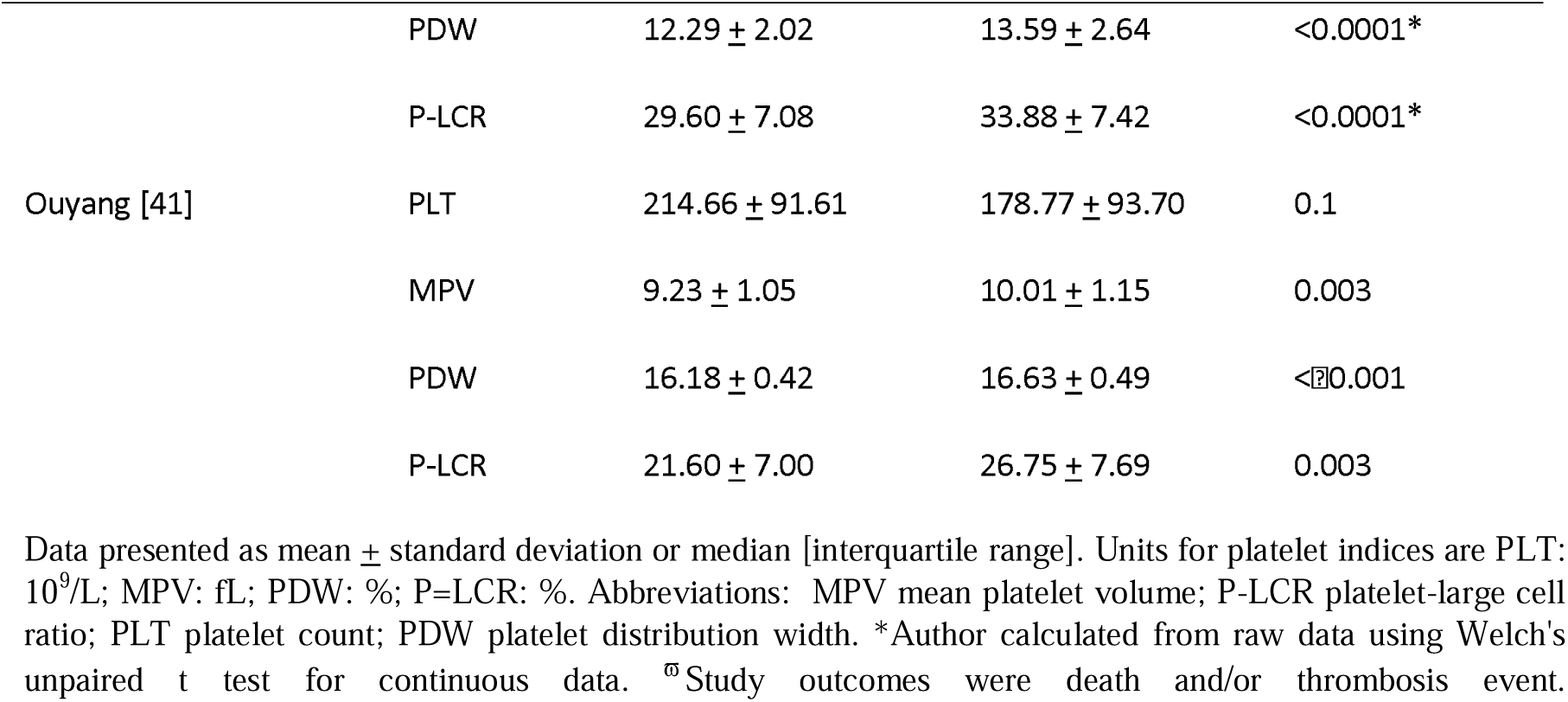
Values of platelet indices in the COVID-19 patient survivor and non-survivor groups

### Quality assessment

Fourteen studies were rated poor and eight were rated fair (Supplementary material 2). Most studies, clearly specified the location, time period and demographics of the selected participants. However, there are several reasons for rating the studies based on potential bias. All studies were cross-sectional in design and the majority were retrospective. Consequently, most authors had no control over the exposure assessment and did not fully describe the methods used to measure the platelet parameters. Only four studies provided the number of eligible patients and the total number included in the study [19, 21, 30, 31]. This inhibited the assessment of participation rate in the other studies. Moreover, no study included a justification for sample size, hence, the authors were unable to make a valid inference about the population being studied. Only three studies clearly described the methods used for the blood counts such as haematology instrument, venepuncture and time/day of test [30-32]. A possible reason for the absence of detail is that data was collected retrospectively from hospital records or other sources, so the authors were unable to identify the method used by the hospital staff to perform the blood counts. Pre-analytical and analytical variables, such as the anticoagulant used, the time between blood collection, storage temperature and instrument type are known to significantly affect MPV measurements [33]. For example, platelets collected into ethylenediaminetetraacetic acid anticoagulant undergo time-dependent platelet swelling and activation. Thus, interpretation of these studies requires a note of caution when considering the reliability of the platelet measurements. In addition to methodology, MPV may be influenced by various demographic factors including age [34, 35]. Of the 22 included studies, six did not statistically compare the age for both non-severe patients or survivors and severe patients or non-survivors [7, 19, 28-30, 36], though it should be noted that two of these studies did statistically compare age between >3 groups [7, 29].

### Platelet size as a predictor of severity in COVID-19 patients

#### Mean platelet volume

Among the 22 studies included in the meta-analysis, 15 reported using MPV in non-severe and severe COVID-19 patients [7, 20, 21, 23, 26-32, 36-39]. Of these, the publication of Alnor et al [30] was excluded as the mean ± SD was unable to be extrapolated due to skewness in the data.

Random-effects meta-analysis revealed a significantly higher mean of MPV levels in severe patients than in non-severe patients (14 studies, SMD = 0.23 [95% CI: 0.07, 0.39], *p* = 0.005, n = 10,382) (Figure 2). There was substantial heterogeneity (*I*2 = 65%) and the funnel plot showed asymmetry, which could be from either heterogeneity of studies or publication bias (Supplemental material 3: Figure 1). Sensitivity analysis based on the time of blood test being taken at hospital admission from five studies [7, 21, 31, 32, 38] gave a slightly higher mean MPV in severe patients (SMD = 0.33 [95% CI: 0.11, 0.55], *p* = 0.004, n = 1551), with a lower heterogeneity (*I*^2^ = 26%). Sensitivity analysis based on ICU admission for six studies [7, 21, 29, 31, 32, 37] did not lower the heterogeneity (*I*^*2*^ = 65%), but the MPV remained significantly higher in the severe group (SMD = 0.40 [0.06, 0.74], *p* = 0.02, n = 1556).

**Figure 2:**
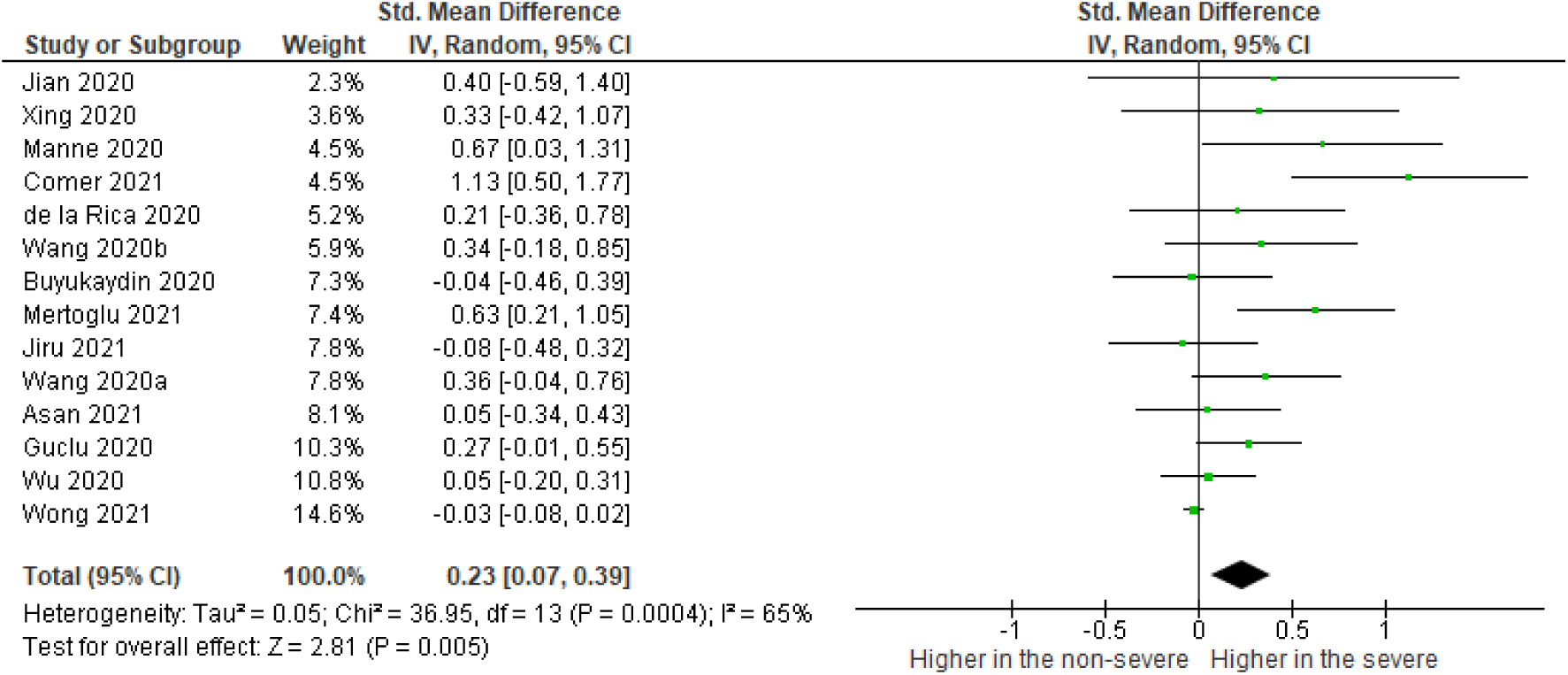
The summary of pooled mean differences of MPV on the observed day after admission between severe and non-severe COVID-19 patients.

A subgroup analysis by quality assessment rating, showed that the MPV was significantly higher in the severe group for both poor- and fair-quality studies, and there was no statistical significance identified between the two subgroups (*p* = 0.99) (Supplementary material 4: Figure 1). However, the heterogeneity for the fair-quality studies was decreased *I*^2^ = 31%), hence, there may be a bias within the poor-quality studies that is causing an increase in heterogeneity. A subgroup analysis for reporting of clear and unclear outcome measures showed no significant difference between the two subgroups (*p* = 0.24) and both demonstrated significantly higher MPV in the severe group (Supplementary material 4: Figure 2). Furthermore, the heterogeneity was *I*^*2*^ = 0% for studies with clearly defined outcome measures, and *I*^*2*^ = 81% for unclear outcome measures.

#### Platelet distribution width

Eight of the 22 included studies reported using PDW in non-severe and severe COVID-19 patients [20-22, 26, 30, 36, 38, 39]. Of these, one study was excluded as the mean + SD was unable to be extrapolated due to skewness in the data [20].

The pooled mean difference of the PDW values for the seven studies was significantly higher in the severe group (SMD = 0.23 [95% CI: 0.07, 0.40], *p* = 0.005, n = 754); and there was no heterogeneity using the random effect model (*I*^2^ = 0%) (Figure 3). Sensitivity analysis showed borderline significance when accounting for time of blood test in three studies [21, 30, 38] (SMD = 0.21, [95% CI: -0.02, 0.44], *p* = 0.07, n = 334, *I*^*2*^ = 0%).

**Figure 3:**
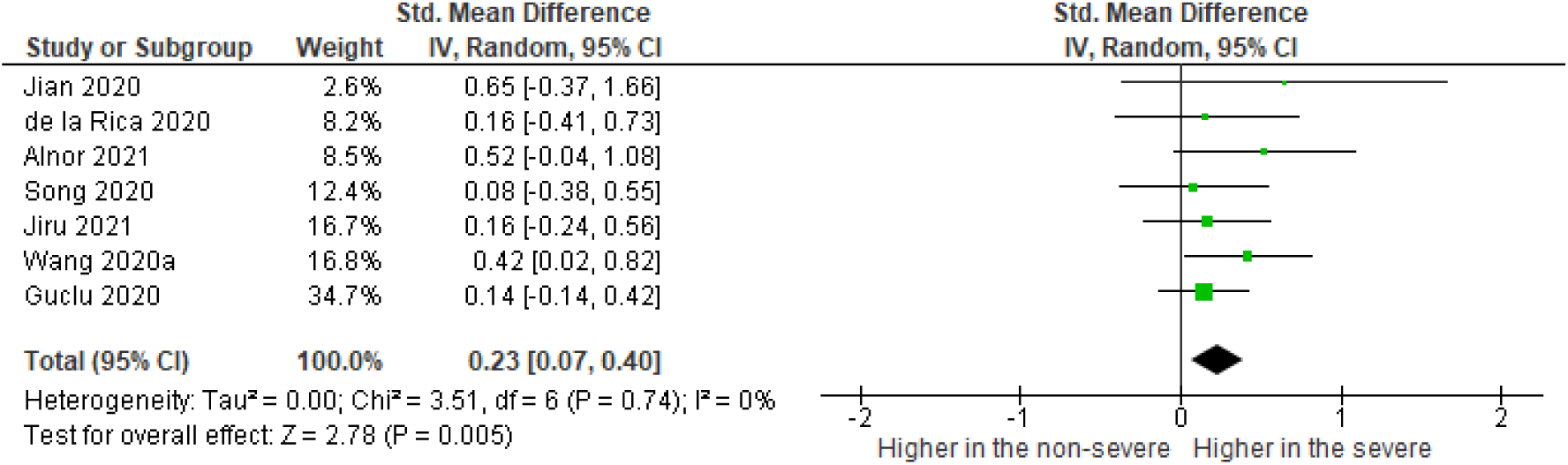
The summary of pooled mean differences of PDW on the observed day after admission between severe and non-severe COVID-19 patients.

#### Platelet- large cell ratio

Four of the 22 included studies reported using P-LCR in non-severe and severe patients [20, 26, 30, 32]. The pooled mean difference of the P-LCR values was higher in the severe group and this result was extremely significant (SMD = 0.60 [95% CI: 0.32, 0.88], *p* <0.0001, n = 1089); however, there was substantial heterogeneity (*I*^2^ = 52%) (Figure 4). A sensitivity analysis based on blood tests taken at admission by excluding Wang et al [26], still showed higher MPV in severe patients (SMD = 0.60, [CI: 0.32, 0.88], *p* = <0.0001, n = 1089) but did not lower the heterogeneity (*I*^*2*^ = 54%).

**Figure 4:**
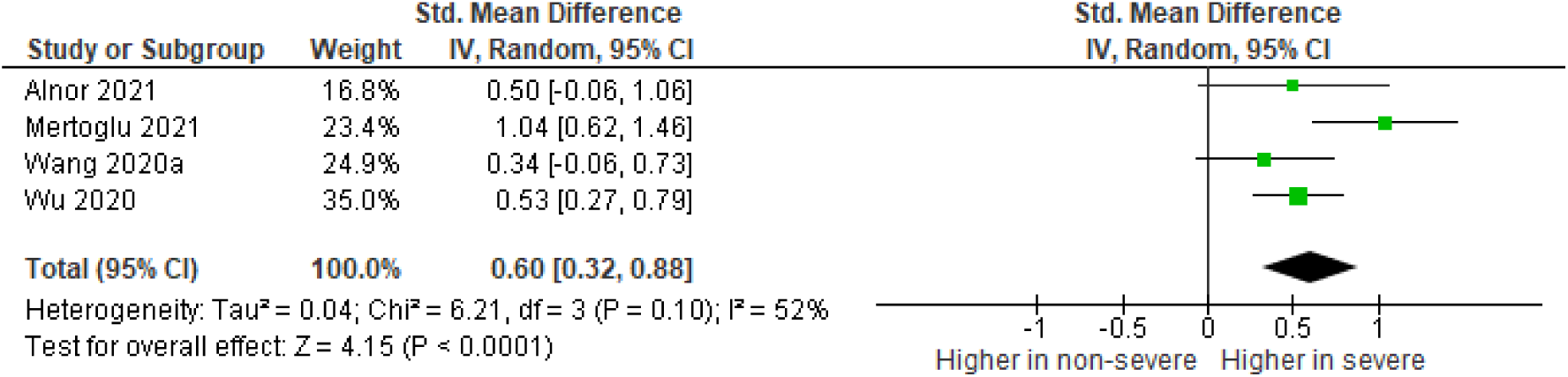
The summary of pooled mean differences of P-LCR on the observed day after admission between non-severe and severe patients.

### Platelet size as a predictor of mortality in COVID-19 patients

#### Mean platelet volume

Eight studies reported using MPV in COVID-19 patients who survived and died [19, 24, 25, 37, 38, 40-42]. All studies measured survival and death, except Barrett et al [25] who measured hospitalised patients who had a thrombotic event and death. The pooled mean difference of the MPV values was significantly higher in the severe group (SMD = 0.34 [95% CI: 0.14, 0.53], *p* = 0.0006, n = 1,337); however, there was substantial heterogeneity by the random effect model (*I*^2^ = 55%) (Supplementary material 5: Figure 1). Sensitivity analysis based on blood tests taken at hospital admission for four studies [19, 24, 25, 38] showed similar results (SMD = 0.40 [95% CI: 0.19, 0.61], p = 0.0002, n = 849) and heterogeneity (*I*^*2*^ = 50%). Similar results and heterogeneity were also shown for sensitivity analysis based on the clinical outcome by excluding Barrett et al [25] (SMD = 0.31 [0.10, 0.52], *p* = 0.004, n = 1237, *I*^*2*^ = 59%).

#### Platelet distribution width

Five studies reported using PDW in COVID-19 patients who survived and died [19, 24, 38, 40, 41]. The pooled mean difference of the PDW values was significantly higher in the non-survivors (SMD = 0.45 [95% CI: 0.19, 0.71], *p* = 0.0007, n = 948); however, there was substantial heterogeneity by the random effect model (*I*^2^ = 68%) (Supplementary material 5: Figure 2). Sensitivity analysis based on blood tests taken at admission for three studies [19, 24, 38] also showed a significantly higher MPV in the severe group (SMD = 0.33 [95% CI: 0.06, 0.60], *p* = 0.02, n = 750), yet conversely, it statistically demonstrated homogeneity (*1*^2^ = 0%).

#### Platelet-large cell ratio

Three studies reported using P-LCR in COVID-19 patients who survived and died [19, 24, 41]. The pooled mean difference of the P-LCR values was higher in the non-survivors and this result was extremely significant (SMD = 0.49 [95% CI: 0.20, 0.77], p = 0.0007, n = 634); however, there was substantial heterogeneity by the random effect model (*I*^2^ = 60%) (Supplementary material 5: Figure 3). Sensitivity analysis based on blood tests taken at admission for two studies [19, 24] also showed a significant overall effect (SMD = 0.42, [95% CI: 0.06, 0.78], *p* = 0.02, n = 536) with substantial heterogeneity (*I*^*2*^ = 75%).

### Probability of higher platelet size in COVID-19 patients

Using the SMD data from five studies [7, 21, 31, 32, 38], the probability (CLES) that a severe COVID-19 patient will have a higher MPV at hospital admission, compared to a non-severe patient, was 59.2% [95% CI: 53.1%, 65.1%] (Figure 3a). By comparison, the probability (CLES) that a severe COVID-19 patient will have higher PDW was 55.9% [95% CI: 50.6%, 62.2%] in three studies [21, 30, 38], and a higher P-LCR, 68.7% [95% CI: 59.8%, 76.5%] in three studies [20, 30, 32] (Figure 5A).

**Figure 5:**
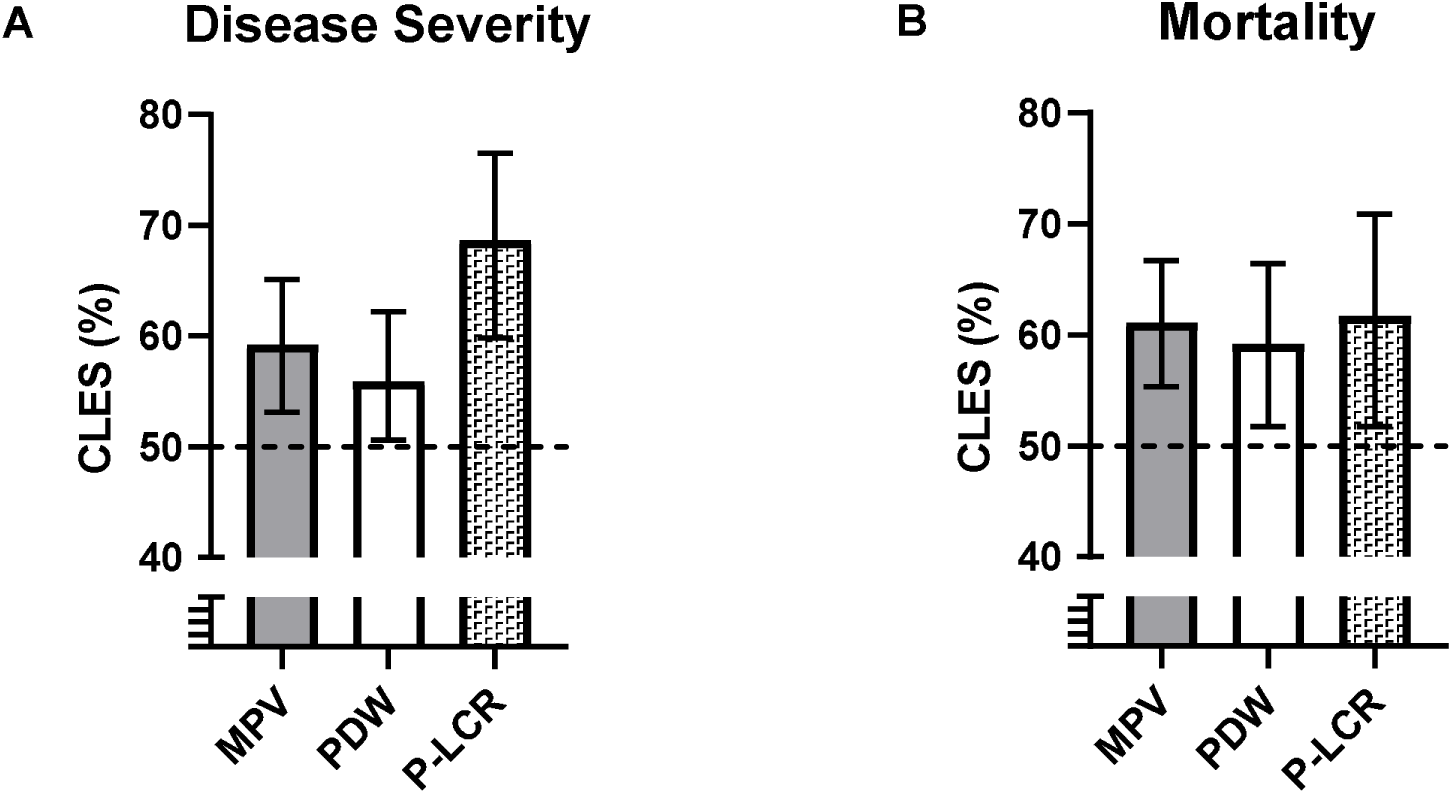
Percentage of severe patients (A) and non-survivors (B) COVID-19 patients that will have higher platelet size than the non-severe or survivors at hospital admission. The dotted line represents the CLES for 50% probability (i.e. no effect). CLES: Common language effect size.

Using SMD data from four studies [19, 24, 25, 38], the CLES for MPV in non-survivors was 61.1% [95% CI: 55.3%, 66.7%]. Indicating that there is a 61.1% probability that a COVID-19 patient who does not survive, will have a higher MPV at hospital admission than those that do survive. By comparison, the probability that a non-survivor will have a higher PDW was similar to MPV (CLES: 59.2% [95% CI: 51.7%, 66.4%] in three studies [19, 24, 38]. A marginally higher P-LCR was also identified (CLES: 61.7% [95% CI: 51.7%, 70.9%] in two studies [19, 24] (Figure 5B).

## DISCUSSION

To our knowledge, this is the first comprehensive systematic review and meta-analysis that assesses platelet size as a predictor for both severity and mortality in COVID-19 patients. There were 22 studies (11,906 patients) and the examination was for the association of MPV, PDW and P-LCR in critically ill and deceased COVID-19 patients. The pooled SMD for all studies of MPV in COVID-19 patients shows that MPV is significantly higher in severe patients.

Subgroup analysis showed that MPV was significantly higher in the severe groups for both poor- and fair-quality studies, and there was a stronger significant association between MPV and severity for studies that reported clearly defined outcome measures compared to studies with unclear outcome measures. These outcomes included the reporting of specific clinical characteristics and using national or local guidance to group patients into severe and non-severe categories. This indicated the importance of reporting clearly described outcomes measures in studies of MPV levels in COVID-19 patients to improve reliability of the meta-analysis findings.

PDW is also associated with severity of disease in the pooled analysis for six studies. However, there was borderline statistical significance in overall effect when accounting for time of the blood test at hospital admission. Nevertheless, the pooled studies for PDW in severe patients gave 0% statistical heterogeneity, implying that there was no variability in the data. Likewise, P-LCR is associated with severity of disease in COVID-19 patients.

Like severity of disease, MPV was significantly associated with mortality in COVID-19 patients. This supports work by Lippi et al [43] who performed an analysis of MPV in pooled data from severe and deceased COVID-19 patients.

More importantly, our study identified a marked increase in the probability of a severe COVID-19 patient presenting with higher P-LCR at hospital admission than a non-severe patient, when compared with MPV and PDW. It also identified that there was a higher probability that a COVID-19 patient who does not survive, will have a higher MPV, PDW and P-LCR at hospital admission than a survivor. Although the three platelet parameters give relatively modest increases for non-survivors.

Most studies included in this review reported a trend for lower PLTs that were insignificantly lower in severe patients and non-survivors. Other studies have reported that patients with severe COVID-19 disease have a PLT only 23 ×10^9^/L to 31 ×10^9^/L lower than those with non-severe disease [44, 45]. The fact that such severely ill patients with systemic immune and coagulation activation maintain reasonable PLTs implies a marked compensatory platelet production response [46].

Elevated MPV is associated with increased risk for thrombotic complications in acute coronary syndrome [14]. High MPV implies an increase in circulating immature platelets and is the body’s response to thrombocytopenia as megakaryocytes respond to increased platelet consumption [46]. Platelet size positively correlates with surface receptor number and ATP content, and large platelets have a higher potential for protein synthesis and bind more fibrinogen [47]. Findings from this study suggest that severe COVID-19 disease is associated with the increased production of larger, younger platelets in response to mild thrombocytopenia. Other studies have reported a trend towards elevated immature platelet fraction in COVID-19 patients [46, 48]. Considering severe COVID-19 is associated with increased numbers of immature platelets in combination with normal PLTs, this could be a mechanism for increased thrombosis in COVID-19 [46].

### Strengths and limitations of the study

Heterogeneity was explored by conducting sensitivity analyses, minimizing the effect of possible confounders. Specifically, the day the blood test was performed and clinical outcome measures. Our data established that these parameters did not influence the results of the meta-analysis for MPV and P-LCR in severe patients, and for all three platelet size biomarkers in non-survivors. A limitation of our study was that most of the included studies were cross-sectional retrospective. Thus, the studies were dependent on the data that was entered into a clinical database and not collected for research from a predesigned protocol. Consequently, some key statistics may not be measured due to unavailable data, and certain variables that have the potential to impact the outcome may not have been recorded at all. For example, only one study excluded patients who had taken platelet medication for >10 days prior to the study [29]. In addition to this, many studies did not describe the full process of the blood collection and analysis as it was conducted by hospital staff prior to the start of the study. This could weaken the conclusions made by the authors. As an example, Wang et al [25] concludes that an MPV-to-lymphocyte ratio of >8.9 was a high risk for COVID-19 severity. However, without a description of the blood test methodology, clinicians cannot determine what laboratory conditions this cut-off would apply to, as different conditions can give varying platelet results (e.g. type of venepuncture tube, haematology instrument or time to analysis).

### Implications for clinical practice and future research

The findings of this meta-analysis provide evidence that MPV and P-LCR are potential biomarkers for prognosis of COVID-19 at hospital admission, with P-LCR being the most important of the two platelet parameters. However, certain aspects remain to be explored to fully demonstrate its use in clinical practice. Future studies should be prospective in design so that researchers can assess multiple outcomes at different time frames. They should give comprehensive methodology which includes careful study design, controlled measurement of platelet parameters, full reporting of how the data were acquired, and appropriate statistical considerations for confounding factors. Moreover, pre-determined cut-off values should be established to investigate the predictive efficacy of these biomarkers.

## Conclusion

MPV, PDW and P-LCR are routinely available in clinical laboratories and are inexpensive tests. The findings of our meta-analysis identified a significant association between MPV and P-LCR at hospital admission with disease severity and mortality, and PDW was significantly associated with mortality. Furthermore, P-LCR is the most important indicator of severe disease in COVID-19 patients at hospital admission, when compared with MPV and PDW. Current evidence is predominantly derived from retrospective design, and future studies are required to accurately determine cut-off values that may be used in the clinical setting.

## Data Availability

Summary data that is not presented in tables, figures or supplemental materials is available from the
authors upon reasonable request.

## Abbreviations

CENTRAL: Cochrane Central Register of Controlled Trials CNKI China Knowledge Resource Integrated
CLES: Common language effect size COVID-19 Coronavirus disease 2019 ICU Intensive care unit
IQR: Interquartile range
MPV: Mean platelet volume
PDW: Platelet distribution width
P-LCR: Platelet-large cell ratio
PLT: Platelet count
PRISMA: Preferred Reporting Items for Systematic Reviews and Meta-Analyses
SARS-CoV-2: Severe acute respiratory syndrome coronavirus 2
SD: Standard deviation
SMD: Standardized mean difference

## Acknowledgements

We thank Jecko Thachil (Manchester Royal Infirmary) for useful discussions at the study development stage and for reviewing the manuscript. We thank Robert Campbell (University of Utah), Xin Chen (Gannan Medical University), Sadia Taj (FMH College of Medicine and Dentistry), Yasemin Ustundag (Bursa Yuksek Ihtisas Training and Research Hospital) and Kenneth Wong (The Chinese University of Hong Kong) for providing data. We also thank Suhail Doi (Qatar University) and Yang Han (The University of Manchester) who provided advise on statistical methodology.

