## Supplementary figures and images for "Platelet size as a predictor for severity and mortality in COVID-19 patients: a systematic review and meta-analysis"

### Supplementary material 3

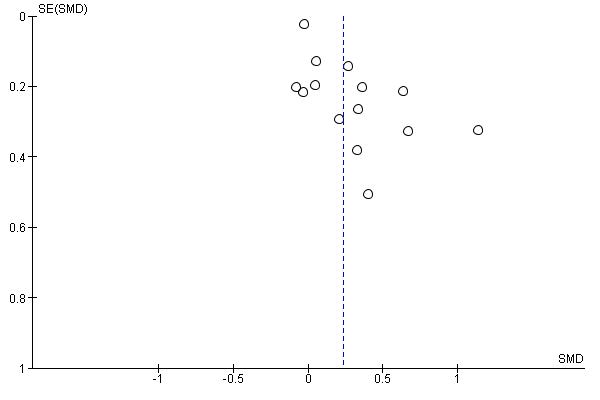

Figure 1: Funnel plot for studies reporting MPV in severe and non-severe COVID-19 patients
