## Supplementary material 4 for "Platelet size as a predictor for severity and mortality in COVID-19 patients: a systematic review and meta-analysis"

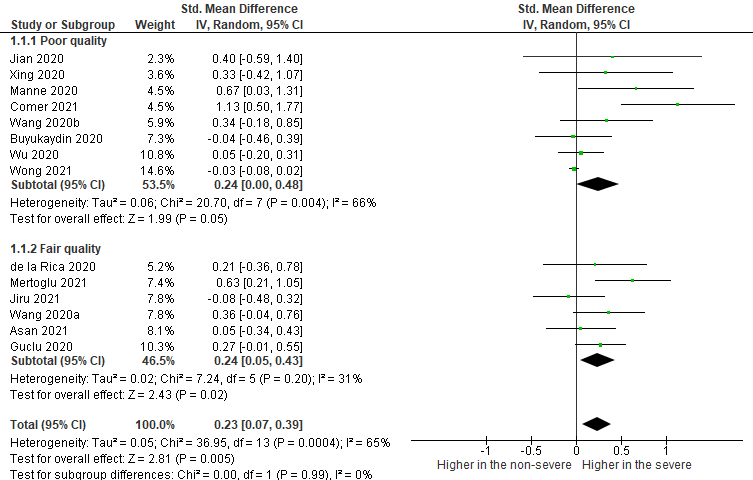

Figure 1: Poor and fair-quality subgroup analysis of the pooled mean differences for MPV in studies of non-severe and severe patients

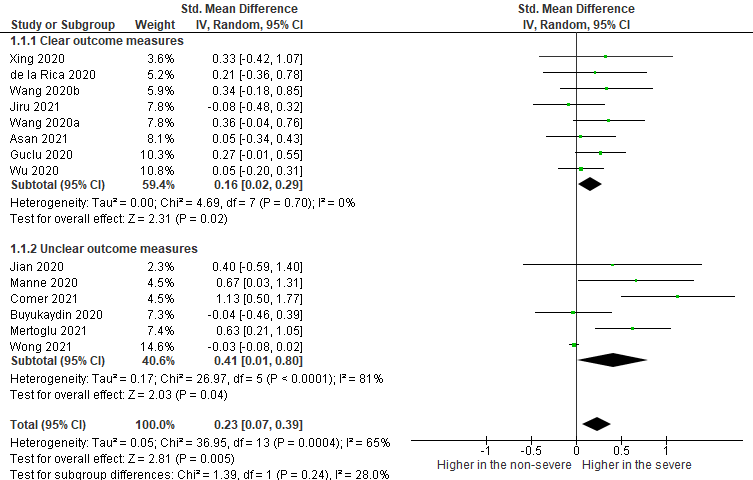

Figure 2: Outcome subgroup analysis of the pooled mean differences for MPV in studies of non-severe and severe patients
