## Supplementary material 5 for "Platelet size as a predictor for severity and mortality in COVID-19 patients: a systematic review and meta-analysis"

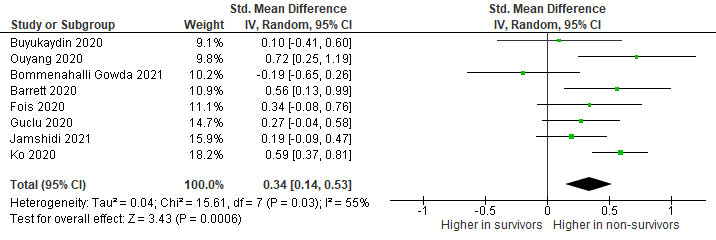

Figure 1: The summary of pooled mean differences of MPV on the observed day after admission between survivors and non-survivors.

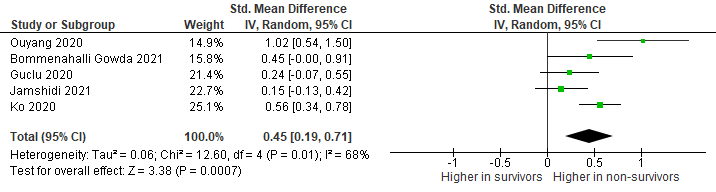

Figure 2: The summary of pooled mean differences of PDW on the observed day after admission between survivors and non-survivors

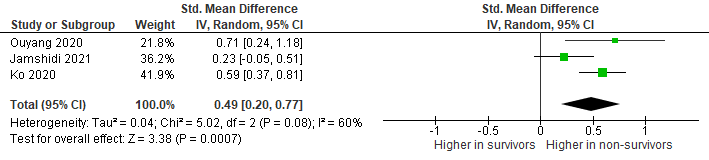

Figure 3: The summary of pooled mean differences of P-LCR on the observed day after admission between survivors and non-survivors
